# A Novel Home-Based Study of Circadian Rhythms: Design, Rationale, and Methods for the Circadia Study

**DOI:** 10.1101/2022.02.25.22271052

**Authors:** Irma Vlasac, Gregory Bormes, Elizabeth Do, Selma Benkhoukha, Naby Diallo, Noah L. Fryou, Siena Gioia, Clarence Joseph, Anne Kuan, Jennifer Lapan, David Oluwadara, the Pepper Team, Richa Saxena, Frank Scheer, Faraji Woodson, Jacqueline Lane

## Abstract

The Circadia Study (Circadia) is a novel “direct to participant” research study investigating the genetics of circadian rhythm disorders. The long-term goals of this study are to better understand the genetics of circadian rhythm disorders, investigate the efficacy and accessibility of an at-home, self-directed DLMO collection, to improve health outcomes in the future for patients with circadian rhythm disorders, and to address the specific needs of the circadian rhythm disorder patient population.

In this manuscript, we briefly outline the standard methods of both circadian biology research protocols and circadian rhythm disorder diagnostic procedures. We describe some of the inherent limitations of current circadian research and diagnostic methods, which motivated our development of and informed the design of the Circadia Study. We discuss the main goals of the Circadia Study, and we outline key features of our study design that build upon current study methods and address limitations. Finally, we describe specific aspects of the Circadia Study, including our study population, data collection methods, and standard operating procedures so that others may replicate aspects of the study design.

The Circadia Study is approved for human subject research by the Mass General Brigham Institutional Review Board, Protocol # 2020P002779.

## Introduction

Circadian rhythm disorders encompass a range of disorders that manifest from both social and environmental factors, such as Shift Work Disorder and Jet Lag Disorder^1^, and from known genetic contributions, such as Advanced Phase Sleep Syndrome (ASPD), Delayed Phase Sleep Syndrome (DSPS), and Non-24 hour Sleep Wake Syndrome (N24)^2^. In this study, we focus on the latter three circadian rhythm disorders, whose genetic links make these circadian rhythm disorders rich with potential for new clinical treatments and diagnostic improvements.

Circadian rhythm disorders are rare diseases, with a prevalence of ASPD at approximately 0.04-0.21% and DSPS at approximately 0.17% across the United States population ^3^,^4^. Patient populations with rare disorders tend to be smaller and more geographically dispersed, creating barriers to studying these populations through traditional study methods, which include recruitment and in-person visits to study locations in metropolitan areas. Additionally, in the circadian rhythm disorder community, the patient population typically lives in a unique temporal niche outside of the 9AM to 5PM societal norms, due to their shifted circadian rhythms, adding an additional barrier to participation in studies of circadian rhythms.

Health disparities stemming from the rare nature of circadian rhythm disorders are compounded first, by a scarcity of geographically dispersed and accessible circadian medicine clinics, and second, by cost of diagnosis for circadian rhythm disorders. Circadian medicine clinics across the United States are few in number and geographically restricted to urban areas, making it difficult for patients outside these areas to make multiple in-person office visits and receive appropriate medical care, diagnosis, and therapies. Additionally, even when patients can travel to a circadian medicine clinic, the cost of diagnosis presents another substantial barrier. Dim Light Melatonin Onset (DLMO), the current gold-standard diagnostic tool in circadian medicine, is not covered by insurance and may incur an out-of-pocket expense of $500-1000.These issues highlight a clear need to increase access to diagnostics, clinical therapeutics, and care for patients living with a circadian rhythm disorder.

Patients may receive a circadian rhythm disorder diagnosis by completing a DLMO at a circadian medicine clinic. However, as previously discussed, the DLMO is not currently covered by insurance, imposing a financial burden on patients seeking a circadian rhythm diagnosis. Alternatively, patients can receive a diagnosis by participating in a circadian rhythm disorder research study that conducts a DLMO and then returns participant data through a diagnostic report. In this case, the cost of the DLMO is offset, but substantial barriers remain. Current studies of circadian rhythms often use traditional methodology for participant recruitment and data collection, which may require in-person visits to circadian/sleep centers for recruitment and/or testing, posing both geographical barriers for accessing circadian/sleep centers and time barriers, due to the long duration of the DLMO collection.

Data collection for circadian rhythm studies that conduct DLMOs typically requires a minimum of 2 visits for patients seeking a DLMO-confirmed circadian rhythm disorder diagnosis. First, at a circadian rhythm research site, the DLMO is conducted. This involves collecting a patient’s salivary melatonin levels on an hourly basis over a period of several hours as a patient remains in a resting position and in dim light conditions. Patient DLMO samples are then processed and analyzed for melatonin levels, and a melatonin phase curve is assessed for variations that indicate a circadian rhythm disorder. For a patient to receive their circadian rhythm disorder diagnosis, they must visit a circadian medicine clinic where a healthcare provider uses the results of the DLMO to confer diagnosis of a circadian rhythm disorder. However, due to the geographically dispersed nature of both circadian rhythm research sites and circadian medicine clinics, the in-person requirements of circadian research present a major obstacle to research participants.

We created the Circadia Study to better understanding the genetics of circadian rhythm disorders, to investigate the efficacy and accessibility of an at-home, self-directed DLMO, to improve health outcomes in the future for patients with circadian rhythm disorders, and to address the specific needs of the circadian rhythm disorder patient population. Here we outline a novel “direct to participant” approach using a completely home-based collection kit and a web-based participant portal as the main interface between study team and both prospective and enrolled participants.

The goals of the Circadia Study are twofold: First, the Circadia Study aims to create an easy-to use toolkit for at-home circadian rhythm assessment through the use of novel in-home based surveys, tests, and collection kits. Through the collection of genetic samples and phenotype assessments with our in-home methodology, Circadia will establish a novel circadian rhythm disorder cohort, providing a foundation for patient-centered scientific resources. These resources may serve for future genetic and related studies as well as possible at-home diagnoses for circadian rhythm disorders via a home-based DLMO kit.

Second, the Circadia Study aims to identify new genetic loci associated with ASPD, DSPS, and N24 through the sequencing and analysis of participant exomes, thereby providing possible loci of interest for targeting in the development of novel therapeutics in the treatment of CRD.

Together, our goals aim to broaden our understanding and elucidate the genetics of ASPD, DSPS, and N24 while increasing accessibility to circadian rhythm disorder diagnostics through self-directed at-home DLMO collections.

### Key Components of the Circadia Study

#### Online Participant Portal

Traditionally, circadian rhythm research studies have required participants to visit a physical location. Either these studies are conducted entirely in-person at a research site^5,6^, or they follow a hybrid model where a participant completes some tasks at a research site and completes others at home with the aid and guidance of a research assistant ^7^. These study models pose significant geographic and time barriers for patients with circadian rhythm disorders. Conducting our research entirely via an online participant portal removes these geographic barriers and cuts down significantly on time commitments for both patients and study team. Through our online portal, study participants are able to consent online and gain immediate access to study material, streamlining our study protocols and making our research study wider-reaching and more inclusive.

The Circadia Study online participant portal was created through collaboration with the Broad Institute Data Science Platform. The portal design is aimed at streamlining all aspects of the study to minimize participant burden and enhance user experience. Prospective participants are directed to the online participant portal through recruitment advertisements and social media. On the public side of the participant portal, anyone may access general information regarding the study, our research, and circadian rhythm disorders. Interested prospective participants will have the opportunity to create an account and log in to access the primary screening questionnaire, a low-burden questionnaire of 11 questions. Participants who are found eligible will then be directed to an online consent form, where they may choose to consent to the study, decline, or return to the main site. The consent form includes graphics explaining the flow of the study and is divided into sections to increase its readability online and to make our study process as clear as possible. A banner providing contact information for study staff is available to participants as they navigate the consent form, should they have any questions and want to contact the study team. Eligible participants who consent to the study are then considered enrolled and have access to the study side of the participant portal, where they may access additional study questionnaires, study instructions, study videos, and other information. The process for a prospective participant, with the use of the online participant portal, may go from recruitment to enrollment in the study within 30 minutes. This can all occur without the participant ever having to contact a study research assistant, thoroughly reducing the time, resources, and participant burden that it would take to enroll a participant in a traditional in-person study. For reference, in a traditional in-person study, a participant would need to schedule a visit with a research assistant, then visit in person with the research assistant to be consented, a process which typically takes one hour, not including travel to and from the study visit site. The Circadia Study at-home protocols are scheduled and completed by the participant based on their personal schedule, accommodating participants who may be active outside of a 9 AM – 5 PM time schedule, with an expected study timeline from enrollment to completion of approximately 4-6 weeks, completed entirely remote within the participant’s home.

#### At-Home DLMO Collection

The current gold standard in circadian medicine to diagnose circadian rhythm disorders is the Dim Light Melatonin Onset (DLMO) assay. In a DLMO assay, hourly salivary melatonin levels are collected in dim light conditions (10 lux) and analyzed to produce a rhythmic profile indicating sleep onset and offset based on the phase curve of melatonin levels^8^. Melatonin is a hormone secreted by the pineal gland and is associated with the endogenous sleep wake cycle in humans. Hourly collections of salivary melatonin samples, when properly collected, allow researchers to assess participants’ naturally occurring sleep-wake cycles and thus detect phase shifts, delays, or advances. The presence of these phase shifts, delays, or advances indicates abnormalities with a patient’s sleep-wake cycle or circadian rhythms^9^.

One of the main challenges with assessing circadian rhythm disorders in patients is the cost and time requirements of a DLMO collection. Traditionally, a DLMO is collected in a sleep lab in dim light conditions (<10 lux) overnight for approximately 8 to 10 hours, with a research assistant guiding participants to collect samples every hour, with analysis of samples costing approximately $12 dollars per sample^10^. Patients are kept in a rested position in dim light conditions for the duration of the collection and then may either be kept overnight or sent home via paid transportation.

The Circadia Study is building on the work of previous circadian studies and is particularly indebted to the work of Helen Burgess^11^, Jeanne Duffy^12^, and Julia Stone^13^. Previous studies have conducted at-home DLMO collections where participants have been required to have in-person contact with the research team to consent to the study and receive study material and instructions for the at-home DLMO collection. Additionally, some studies have had research assistants visit study participants in their homes to prepare a DLMO collection space and review collection instructions prior to the participant collecting their samples independently. Concerns regarding at-home DLMO saliva sampling from the previous studies include a) ensuring participants are in sufficiently dim light to maintain and not suppress endogenous melatonin levels through bright light and b) ensuring participant compliance with sample timing. To address the concerns of sufficient dim light for the DLMO saliva sampling, the Circadia Study designed our at-home kit to provide participants with appropriate material to create dim light conditions within a chosen collection space in a participant’s home. This includes material to block out light from windows, and 16 previously validated tea lights that, when placed in a 4×4 grid, measure at approximately 3 lux (0.1 melanopic lux), ensuring participants are within dim light conditions. To further ensure appropriate dim light conditions are met and maintained, a lux meter and instructions for use are provided to participants to measure their dim light condition space prior to beginning their DLMO collection. Additionally, light exposure data is captured through a light sensor on the actigraphy watch, which participants are requested to wear for the duration of the at-home study protocol. To address concerns regarding participant compliance, Circadia is using both SMS text messages and the phone application Galarm, which allows the study team to send alarms to a participant’s study smartphone to remind participants to collect samples at the appropriate times during the DLMO collection. Participants will also use a Medication Electronic Monitoring System (MEMS) cap, which is a bottle lid traditionally used for monitoring compliance with medication but adapted for the study to monitor the times at which participants open their collection bottles to remove a collection salivette, further providing an objective measure of compliance with study protocol. The design of the Circadia Study is focused on minimizing participant burden and study cost. We provide written and visual instructions and all study materials necessary for participants to complete their at-home sample collections, thoroughly reducing the need for in-person resources and research assistant working hours.

The Circadia Study has created a suite of resources for at-home circadian assessment, which includes both physical materials provided via the at-home kit and online instructional videos, checklists, and written guides to aid participants during their study participation. The material provided for at-home circadian assessment may be found in Table 1, which specifies the motivation for providing each material and whether the material is hosted online through the participant portal or provided in the at-home kit.

**Table 1:** Circadia Study Participant Resources

| Location | Item | Purpose |
| --- | --- | --- |
| Circadia Participant Portal | Attestations | Confirm participation and understanding of study activities |
|  | Instructions | Provide online and downloadable instructions of study activities |
|  | Unboxing Video | Provide visual guidance of at-home kit components |
|  | DLMO Instructional Video | Provide visual guidance of DLMO sample collections using at-home kit |
|  | Checklist | Outline steps of study protocol compliance for participants |
| Circadia At-Home Kit | Study smartphone | Galarm application: schedule alarms at sample collection times<br>SMS: send reminders for sample collection times and to offer guidance<br>Web Browser: Access to instructions and guidance via the participant portal |
|  | At-Home Kit Instruction Packet | A physical copy of the instruction packet is provided to aid participants with their At-Home Kit; a digital copy of these instructions is available on the participant portal |
|  | Actiwatch | Participants are asked to wear an Actiwatch continuously for ~4 weeks. This Actiwatch data will supplement participants' self-reported sleep logging data |
|  | Aluminum Foil, black plastic bags, and painter's tape | Used to block out lights from any windows within collection space |
|  | Blue light blocking glasses | Used for when a participant leaves their collection space and may possibly be exposed to blue light |
|  | 18 tea lights | 16 tea lights provided to be set up in a 4x4 grid to provide dim light conditions (>10 lux) |
|  | MEMs Adherence cap and bottle labeled for DLMO Collection #1 containing (9x) cotton swabs | MEMs adherence cap tracks each time the bottle is opened for a salivette to be retrieved, allowing for validation of timing by study team post collection |
|  | Capped bottle labeled for DLMO Collection #2 containing (9x) cotton swabs | An additional bottle is provided for the second DLMO collection, to be replaced with the MEMs Adherence cap used during the first collection |
|  | 18 salivettes labeled for each saliva collection in DLMO #1 and DLMO #2 | Salivettes are pre-labeled to minimize participant error during sample collection |
|  | Extra sticker labels, extra salivettes, and sharpie | Participants will be given extra labels, salivettes, and a Sharpie marker to label their own tubes in case one of the pre-labeled tubes malfunctions |
|  | Extra soft toothbrushes, tylenol, and medical tape | Miscellaneous items to aid participants in their DLMO collections and actigraphy |
|  | Silver insulated envelopes labeled for DLMO #1 and DLMO #2 | Insulated envelopes help keep the saliva samples at a low temperature during return shipment |
|  | Return shipping label, return shipping pouch, and strips of packing tape | Participants will be given all the materials they need to return ship their kit to the study team |

#### At-Home Kit

The Circadia Study created an at-home kit to be used in conjunction with the participant portal, to guide participants through the study protocol and ensure appropriate sample and data collection.

The at-home kit was specifically created and designed to provide for all participant study needs for the duration of the at-home portion of the study. In an effort to minimize participant burden, the Circadia Study essentially set out to create a sleep lab within someone’s home, designed specifically for the collection of the DLMO samples. We designed the kit with a focus on accessibility and inclusivity; participants will be provided with everything needed for successful completion of the study protocol, including a smartphone device to allow participants to access the online components of the study. For participants without access to a secure internet connection, a smartphone will be provided with a data plan to be used for the duration of the study in regions that have data access.

The following materials will be provided in the at-home kit:

- Study smartphone preloaded with study applications and bookmarked, relevant websites
- Actiwatch (Wrist activity monitor with light sensor)
- DLMO test kit for 2 sets of collections
- Aluminum foil for dim light environment
- Bottle with time tracker lid
- Light Meter

Additional miscellaneous material, such as toothbrushes, night lights, labels, and markers will be provided within the kit as well, to ensure all materials required to complete the study protocol are provided to the participant.

### Study Design

The Circadia Study is a cohort study aimed at recruiting approximately 200 participants diagnosed with circadian rhythm disorders. Of the 200 participants with circadian rhythm disorders, we expect our primary recruitment efforts will include approximately 75 participants with ASPD and 115 with DSPS, and we will eventually expand our recruitment efforts to include 10 participants with sighted N24. Recruitment for the Circadia Study will exclude the following participants, due to potential non-genetic impacts on schedule and endogenous circadian rhythms: those who are aged 22 years and younger and/or are enrolled in college^14^; pregnant participants or participants who have given birth in the past year^15^; individuals taking medications that inhibit or interfere with melatonin production; individuals who have suffered from a traumatic brain injury, stroke, or live with a seizure disorder^16^; individuals with gingivitis, xerostomia, or periodontitis due to the nature of the DLMO collection and potential to exacerbate these oral conditions; and individuals who cannot read and write in English at an 8th grade level, since the at-home protocol requires multiple pages of reading instructions and completing questionnaires. A future goal of the Circadia Study is to produce our study material in multiple languages in line with our focus on study accessibility.

A comprehensive description of the methods used in the Circadia Study may be found in the *Circadia Study Standard Operating Procedure*, available as supplementary material and as a downloadable resource on the Circadia study website. The overall approach to the Circadia Study is focused on minimizing participant burden to entry to the study, followed by streamlined, direct to participant access to study material and study collection. With the use of an online participant portal specific to the Circadia Study, participants may learn about the study, complete an eligibility instrument, and enroll in the study through the participant portal. Once enrolled in the study, participants begin completing required study material online, which includes scheduling at-home DLMO sample collections and sleep logging. Once a participant has completed their scheduling and at least one week’s worth of sleep logging, a participant will receive an At-Home kit in the mail. The At-Home Kit contains a study smartphone where participants may continue sleep logging as well as access the participant portal and other study applications, such as Bitesnap for food logging, begin immediate activity monitoring through an Actiwatch, and access an unboxing video, where participants are guided through all study materials provided in the at-home kit. Using provided study materials and both printed and video guidance, participants will be able to create dim light conditions within their home to collect their DLMO samples on two different days approximately one week apart. The study team will use the two sets of collected DLMO samples to validate the participants’ self-reported diagnosis in conjunction with sleep-wake logging data and actigraphy data. A participant’s overall participation in the study is expected to take approximately 4-6 weeks, starting from enrollment through end of study sample return.

### Population

Across the United States, the prevalence of DSPS is approximately 0.17% and ASPD is approximately 0.04-0.21%^17^. While N24 is observed in approximately half of the non-sighted population, individuals with sighted N24 is much more rare^18, 19^.

### Data Acquisition

#### Questionnaires

Participants in the Circadia Study will have a total of 18 study questionnaires that will be completed over the course of the participant’s study duration. Study questionnaires chosen for participant completion focus on circadian phenotyping, sleep assessments, emotional-behavioral assessments, and sociodemographic information. The Circadia Study is using both previously validated questionnaires and new questionnaires to gain insight into our cohort. The 18 questionnaires may be separated into 4 categories: Circadian Type/Phototype questionnaires; Sleep questionnaires; Morning-Evening Preference questionnaires; and Emotional/Behavioral questionnaires, as shown in Table 2.

**Table 2:** Circadia Study Participant Questionnaires

| Category | Instrument | PMID |
| --- | --- | --- |
| Circadian Type | Circadian Type Inventory | 8440215 |
|  | Circadian Phototype Questionnaire |  |
| Sleep | Epworth Sleepiness Scale | 1798888 |
|  | Insomnia Severity Index | 21532953 |
|  | Pittsburgh Sleep Quality Index | 2748771 |
|  | Restless Legs Syndrome questionnaire | 25023924 |
|  | STOP BANG Sleep Apnea Questionnaire | 26378880 |
|  | Sleep Hygiene Questionnaire | 9790493 |
|  | Dreaming Questionnaire |  |
| Morning Evening Preference | Morning Evening Questionnaire | 1027738 |
|  | Munich Chronotype Questionnaire | 12568247 |
|  | Sleep, Circadian Rhythms, and Mood (SCRAM) Questionnaire | 30395721 |
| Emotional/Behavioural | Sociodemographic and Lifestyle Questionnaire |  |
|  | Beck Depression Inventory | 13688369 |
|  | Seasonal Affective Disorder (SAD) Questionnaire |  |
|  | Generalized Anxiety Disorder (GAD-7) Questionnaire | 16717171 |
|  | Emotional Eater Questionnaire | 22732995 |

#### Home DLMO Collection

Participants are instructed to self-schedule two at-home DLMO collections after completing one week’s worth of sleep logging. One week’s worth of sleep logging is required prior to scheduling DLMO collections so the study team can determine the timing of hourly DLMO salivary sample collection^20^. Participants are asked to schedule their two DLMO collections (DLMO Collection 1 and DLMO Collection 2) approximately one week apart from each collection using an online booking tool, providing at least two weeks from their enrollment date to their first scheduled DLMO collection. This allows sufficient time for sleep logging and for shipment of the Circadia At-Home Kit. Self-scheduling reduces participant burden by allowing participants to decide on a DLMO collection schedule that works with their personal life and work schedules.

The Circadia At-Home kit provides detailed instructions and all required material to create dim light conditions within the participant’s home. Dim light conditions while collecting melatonin salivary samples are essential to capture the participant’s endogenous melatonin profile, since higher light intensity reduces melatonin levels^21^. To ensure participants successfully create a dim light collection space (<10 lux), participants are provided with 18 tea lights, 16 for use within their dim light collection space and two additional tea lights to be used in a restroom, should a participant need to use one during the study protocol. Measured lux from the 16 tea lights, when placed in a 4×4 grid and measured approximately 3 feet away reads at approximately 3 lux, well below the 10 lux dim light threshold. Melanopic lux is a metric used to describe non-visual light effects on the eye. The light emitting from the 4×4 grid comprised of 16 tea lights measures below 1 melanopic lux, indicating that participants melatonin levels will not be affected by possible blue light exposure from the provided study tea lights^22^. Participants are instructed to use a provided light meter within the At-Home Kit to measure light levels, in lux, once they have set up their dim light collection space. This allows participants to know whether they need to further prepare their space to achieve dimmer light settings. Once they have achieved dim light conditions in compliance with study protocol, they may begin sample collection. To further ensure participants maintain dim light conditions and to minimize possible risks of exposure to higher intensity light, participants are provided with blue light blocking glasses and instructed to wear them in any instances where they would need to leave the collection space (e.g. to go to the bathroom) and may be exposed to light levels greater than 10 lux. These protocols ensure the accurate capture of endogenous melatonin levels during sampling^23^.

Additionally, the At-Home Kit is created to be used with online portal material and video tutorials with captions, ensuring participants have sufficient guidance to execute the self-directed DLMO collection study protocol. Participants will be guided through an instructional video for unboxing of their At-Home Kit to familiarize themselves with all components of the At-Home Kit upon its receipt. A quick response (QR) code on the outside of the At-Home Kit directs participants directly to the Circadia unboxing video for participants with access to a QR capable device, such as a smart device. The At-Home Kit contains checklists and printed guides that are also available in the participant portal to provide further guidance to participants as they complete the study protocol. To guide them through the DLMO collection, participants can watch a step-by-step instructional video demonstrating how to prepare a collection space to achieve dim light conditions, how to collect hourly melatonin saliva samples, and how to store samples after completion of each sample collection. To further guide participants and to aid the study team in validating participant study protocol compliance, study protocol attestations are expected from participants at various points within the at-home protocol, including prior to beginning each DLMO collection to confirm that participants have followed study protocol preparations and created a dim light collection space, and after the collections to confirm that samples are properly stored and shipped.

#### At-Home Actigraphy Collection

To support the self-reported sleep logging data, participants perform home actigraphy collection using an Actiwatch, a watch-like device that is worn on the wrist. Participants are instructed to wear the Actiwatch on their non-dominant wrist for the duration of the at-home protocol; this will be for 24 hours a day for about 4 weeks. The Actiwatch helps track sleep and wake times by measuring light levels in participant’s environment.

Additionally, participants are instructed to press a button on the Actiwatch each time they go to sleep and wake up. Actiwatch data is analyzed using Actiware software and manually scored. This actigraphy data is essential to fill in missing gaps in the sleep logs and otherwise validate sleep logging data.

#### At-Home DNA Sample Collection

In addition to saliva samples collected during the dim light melatonin assay, study participants provide a single saliva sample for exome sequencing. DNA samples are collected through a diagnostic kit sent third-party company GBF, Inc. Genetic diagnostic kits will be shipped concurrently with the Circadia Study At-Home Kit, and both kits will be returned together. Genetic data collected during the *At-Home DNA Sample Collection* will be used to sequence participant exome sequencing and analysis, from which data we hope to identify new genetic loci associated with ASPD, DSPS, and N24.

#### Optional Protocol: Blood Draw

Since we are piloting the first completely remote collection of saliva-based DLMO assays, we plan to corroborate results of the saliva-based DLMO assays through an optional protocol where participants may consent to providing an optional blood sample as part of the study protocol. This would require an in-person visit on the morning of either day of a participant’s scheduled DLMO collection. Participants local to the study site may choose to participate in the optional study protocol, but it is not required for study participation or completion. The timing of the blood draw will be decided based on a participant’s circadian rhythm disorder diagnosis as follows:

> Advanced Sleep Phase Syndrome: Between 6 AM and 10 AM
>
> Delayed Sleep Phase Syndrome: Between 10 AM and 2 PM

A total of 8 milliliters of blood will be drawn intravenously by qualified personnel and then processed and analyzed for melatonin concentration levels.

## Discussion

Patients living in the United States with circadian rhythm disorders face multiple barriers to circadian medicine and circadian research, such as geographic barriers due to the limited number of circadian medicine clinics in the United States, financial burdens of diagnostics, and accessibility of circadian research sites and protocols that may provide an alternate route of diagnosis. Specifically, the circadian rhythm disorders ASPD, DSPS, and N24 have known genetic contributions, but barriers to circadian research and time-consuming in-person study protocols add additional difficulties to participants in circadian research studies. Circadian studies that allow for in-home protocols and sample and data collection are necessary to reduce participant burden and improve accessibility and inclusion in circadian rhythm research. These studies provide a path for the circadian community to better study the genetics of circadian rhythm disorders, benefiting those living with circadian rhythm disorders through improved diagnostics and therapeutics.

The Circadia Study is the first fully remote clinical circadian research study aimed at elucidating the genetics of ASPD, DSPS and N24, using novel and innovative approaches to ease patient burden while keeping accessibility and inclusivity at the forefront of our research design. Using a ‘direct to participant’ approach, the Circadia Study has created a study where participants may complete the entire study process, from recruitment to completion of study protocol, all from within their home with an expected study duration of 4-6 weeks. Participants are provided with all material needed to essentially create a sleep lab within their home and are only expected to use their own personal electronic devices for the duration of consent and enrollment, prior to a study kit being mailed to their homes.

Traditional circadian research methods rely on in-person recruitment and enrollment efforts. Recent studies have begun using in-home collection techniques but require the in-person guidance of a research assistant, which still limits the geographic capacity of the study and places a sizeable burden on participants to accommodate the study schedule. While traditional study methods provide for greater control of study conditions and compliance, they also pose limitations to participants and may reduce the diversity of participants seeking to engage in circadian rhythm research studies.

We expect the results of the Circadia Study to inform both current circadian research methodology and the practice of circadian medicine. Adapting a direct to participant approach for circadian research may increase the diversity of patient populations that may participant in circadian research, extending research outside of the lab to be far reaching. This is especially important in the context of rare disease, where communities may be smaller and more geographically dispersed. Through our study findings, we hope to establish a validated at-home DLMO that in the future may be adapted to clinical use, wherein clinicians may order an at-home DLMO for patients seeking a diagnosis for a circadian rhythm disorder. With the reduced costs of the at-home DLMO, it may be feasible for the at-home DLMO to be covered by insurance or, at the very least, more accessible in cost if it must be paid for out-of-pocket. We hope to establish the Circadia Study as a resource for the circadian research community, providing our study design and material as guidance for those seeking to adapt similar ‘direct to participant’ methodology to their own studies, furthering the science of circadian rhythms and offering greater support to the circadian rhythm disorder community.

## Data Availability

All data produced in the present study are available upon reasonable request to the authors

## Footnotes

1 Sack et al., “Circadian Rhythm Sleep Disorders,” November 1, 2007.

2 Sack et al., “Circadian Rhythm Sleep Disorders,” November 2007.

3 Curtis et al., “Extreme Morning Chronotypes Are Often Familial and Not Exceedingly Rare.”

4 Ancoli-Israel et al., “A Pedigree of One Family with Delayed Sleep Phase Syndrome.”

5 Lewy, Cutler, and Sack, “The Endogenous Melatonin Profile as a Marker for Circadian Phase Position.”

6 Klerman et al., “Comparisons of the Variability of Three Markers of the Human Circadian Pacemaker.”

7 Burgess et al., “Home Circadian Phase Assessments with Measures of Compliance Yield Accurate Dim Light Melatonin Onsets.”

8 Lewy, Cutler, and Sack, “The Endogenous Melatonin Profile as a Marker for Circadian Phase Position.”

9 Pandi-Perumal et al., “Dim Light Melatonin Onset (DLMO).”

10 Molina and Burgess, “Calculating the Dim Light Melatonin Onset.”

11 Burgess et al., “Home Dim Light Melatonin Onsets with Measures of Compliance in Delayed Sleep Phase Disorder.”

12 Pullman, Roepke, and Duffy, “Laboratory Validation of an In-Home Method for Assessing Circadian Phase Using the Dim Light Melatonin Onset (DLMO).”

13 Stone et al., “The Role of Light Sensitivity and Intrinsic Circadian Period in Predicting Individual Circadian Timing.”

14 Crowley et al., “A Longitudinal Assessment of Sleep Timing, Circadian Phase, and Phase Angle of Entrainment across Human Adolescence.”

15 Martin-Fairey et al., “Pregnancy Induces an Earlier Chronotype in Both Mice and Women.”

16 Castriotta et al., “Prevalence and Consequences of Sleep Disorders in Traumatic Brain Injury.”

17 Schrader, Bovim, and Sand, “The Prevalence of Delayed and Advanced Sleep Phase Syndromes.”

18 Sack et al., “Circadian Rhythm Abnormalities in Totally Blind People.”

19 Malkani et al., “Diagnostic and Treatment Challenges of Sighted Non–24-Hour Sleep-Wake Disorder.”

20 Burgess and Eastman, “The Dim Light Melatonin Onset Following Fixed and Free Sleep Schedules.”

21 Rahman et al., “Characterizing the Temporal Dynamics of Melatonin and Cortisol Changes in Response to Nocturnal Light Exposure.”

22 Nowozin et al., “Applying Melanopic Lux to Measure Biological Light Effects on Melatonin Suppression and Subjective Sleepiness.”

23 Sasseville et al., “Blue Blocker Glasses Impede the Capacity of Bright Light to Suppress Melatonin Production.”

